# A multistate model of frailty progression after severe infections in adults ≥65 years in England: a matched-cohort study

**DOI:** 10.64898/2026.06.16.26355787

**Authors:** Kwabena Asare, Kathryn E. Mansfield, Georgia R. Gore-Langton, Eleanor Barry, Ruth Keogh, Vincent Lo Re, Maria C. Rodriguez-Barradas, Amy C. Justice, Christopher T. Rentsch, Charlotte Warren-Gash

## Abstract

**Background:** Evidence on frailty progression following severe infections is limited. We compared rates of transition to greater frailty or death between adults with and without severe infection in England.

**Methods:** We conducted a matched-cohort study among adults aged ≥65 years (1,452,117: median age 76 years, 45% male) in Clinical Practice Research Datalink Aurum (2006-2019). Adults with severe infection (hospitalised primarily due to infection) were matched on calendar time to individuals without severe infection on age, sex, and primary care practice. The admission date was used as index date and same was assigned to matched unexposed adults. We measured frailty using Electronic Frailty Index, a proportion of 36 health deficits in validated categories (Fit 0-0.12, Mild >0.12-0.24, Moderate >0.24-0.36, Severe >0.36).

In a time-varying Markov multistate model, we focused on forward transitions from baseline or intermediate frailty states to higher states or death. For each transition, we used Cox regression to estimate cause-specific transition hazard ratios (HR) with 95% confidence intervals (CIs), comparing adults with and without severe infection. We adjusted for baseline frailty score, age, sex, deprivation, harmful alcohol use, smoking, and primary care infection history 5 years before index date. We estimated state occupancy probabilities, and expected length of stay (ELOS) in each state at year five among adults with and without severe infection. We explored effect modification by infection type.

**Results:** Across all transitions, severe infection was associated with higher adjusted hazards of transitioning to worsening frailty or death, HR, 95% CI: (**fit to**: mild[1.56, 1.54-1.58], moderate[2.51, 1.79-3.51], death[4.57, 4.50-4.65]; **mild to**: moderate[1.52, 1.50-1.53], severe[1.90, 1.43-2.52], death[2.67, 2.64-2.70]; **moderate to**: severe[1.40, 1.38-1.42], death[1.87, 1.85-1.90]; **severe to** death[1.48, 1.46-1.50]). Transition hazard ratios were strongest for lower respiratory tract infections, followed by sepsis, urinary tract infections, meningitis/encephalitis, gastroenteritis, and skin and soft tissue infections. At five years, adults with severe infection had higher probabilities of transitioning to greater frailty or death across all transitions and lower ELOS in each frailty state than those without severe infection.

**Interpretation:** Severe infections may accelerate frailty deterioration in older age. Prevention through vaccination, early detection, and prompt management may help mitigate this decline.

**Research in context:** *Evidence before this study:* We searched PubMed (inception to May 08, 2026), for published articles evaluating the association between infections and frailty progression with multistate modelling including studies that focused on one transition only with no language restrictions. We used the search terms [(infection OR infectious) AND (frailty OR frail) AND optional (progression OR multistate OR multi-state OR transition)]. We identified five longitudinal studies that compared transition hazard rates from a baseline fit state to pre-frailty or frailty during follow-up between individuals with specific infections (SARS-CoV-2, HIV, history of 20 infections based on International Classification of Diseases [ICD] codes and prescribed medications, cytomegalovirus antibody concentration quartiles, and seropositivity of herpes viruses [HSV 1 and 2, varicella-zoster, and Epstein-Barr]) versus those without infection. The studies found that infection or infection history increased the hazards of transition from a fit/pre-frail state to frailty after adjusting for confounders. The hazards of frailty recovery from frail to fit state was higher among individuals with HIV infection than those without. None of the studies accounted for the competing risk of death, and all modelled transitions along a single pathway between fit/pre-frail and frail states. Furthermore, frailty was conceptualised as a binary outcome, whereas validated frailty risk prediction models typically define frailty in progressive categories of deterioration, commonly classified as fit, mild, moderate, and severe. There is limited evidence on the risk of transition from baseline and intermediate frailty states to more advanced states following infection, while accounting for competing risk of death. To identify subgroups for targeted care, it is important to determine whether frailty deterioration after severe infection varies by infection type (sepsis, urinary tract infection, skin and soft tissue infection, meningitis/encephalitis, lower respiratory tract infection, and gastroenteritis), age, sex, social deprivation, diabetes, and dementia.

*Added value of this study:* Our study compared rates of transition to greater frailty or death between older adults with and without severe infection (hospitalisation primarily for infection). We found that severe infection increased the hazards of transition from baseline and intermediate frailty states to more advanced frailty states or death. From each same starting frailty state, outgoing transition hazards increased progressively towards more advanced frailty states or death. The hazards of progression to greater frailty or death also varied by infection type, with the strongest associations observed for LRTI, followed by sepsis, UTI, meningitis/encephalitis, gastroenteritis, and skin and soft tissue infections.

*Implications of all the available evidence:* Our findings underscore the importance of infection prevention in reducing the risk of frailty progression in older age. Additional studies are required to explore other wider life-course influences on frailty, to guide the development of comprehensive preventive strategies.

## Introduction

Frailty reflects diminished physiological reserves and increased vulnerability to adverse outcomes after health stressors.^1^ It is strongly associated with loss of an individual’s ability to perform physical and cognitive functions central to their personal quality of life.^2,3^ While frailty is common with advanced age, some ageing adults remain robust while others experience early signs of frailty and may rapidly progress.^1^ This variation suggests that preventable life course factors including acute health events may alter later-life frailty trajectories and contribute to differences in ageing outcomes leading to poor quality of life and early mortality.^1,4^ As longevity increases globally, identifying preventable and/or modifiable risk factors for frailty is increasingly becoming important to inform interventions to prevent frailty and support healthy ageing.^2,3^

Severe infections may exacerbate frailty trajectories through physiological decline, reduced physical function, and possibly accelerated dementia.^1,5–12^ Evidence of specific infections driving frailty progression is emerging, with many studies showing high frailty at infection onset.^5,6,13–23^ Fewer studies have demonstrated progression of frailty after infection although results may be biased by competing risk of death.^13,14,24^ Likewise, small differences in continuous frailty scores reported by some studies can be difficult to interpret in terms of clinical significance.^13^ Furthermore, frailty is often treated as a binary outcome but as it exists on a continuum, it is more meaningfully represented as ordered categories of progression.^25,26^ Thus, understanding risk of transitioning between multiple frailty states, while incorporating the competing risk of death, may improve our understanding of how severe infections influence frailty trajectories.

We hypothesised that severe infections would accelerate transitions to greater frailty, while accounting for the competing risk of death. We used a multistate model in a matched-cohort study of older adults (≥65 years) in England to examine whether severe infection, defined as hospitalisation primarily due to infection, was associated with progression to greater frailty or death.

## Methods

### Study design, population, and data sources

We conducted a matched-cohort study using routinely collected electronic health records from adults aged ≥65 years who received primary health care in practices contributing to England’s Clinical Practice Research Datalink (CPRD) Aurum (March 2025 data release) between 01 April 2006 to 31 December 2019 (to exclude the impact of the COVID-19 pandemic on healthcare access and service delivery). CPRD Aurum contains pseudonymised primary care data representing approximately 30% of the population in England and is broadly representative with respect to age, sex, ethnicity, and geographic region.^27,28^ We additionally used linked datasets: (1) Hospital Episode Statistics Admitted Patient Care (HES-APC) to identify severe infection events;^29^ and 2) Index of Multiple Deprivation (IMD) to assess area level socio economic deprivation.^27^

Participants became eligible for inclusion from the latest of the study start date (01 April 2006), their 65th birthday, or one year after registration with the primary care practice (to ensure adequate ascertainment of baseline health status). Individuals with a severe infection (hospitalised primarily due to infection), were matched to a maximum of five individuals without a recorded severe infection. Matching was performed without replacement, on calendar time, age, sex, and primary care practice. Follow-up commenced at index date and continued until the earliest of study end date (31 December 2019), death, last data collection from practice, and among comparators only, the occurrence of the first severe infection, at which point they were reclassified as exposed and matched to a comparator.

### Exposure

Severe infection was defined as the first hospital admission after age 65 years with infection recorded as the primary diagnosis. To identify this, we used the ICD-10-coded infection recorded in the first diagnostic position of any episode within that hospital admission. In England, hospital admissions may consist of one or more episodes corresponding to periods of care under different consultants. We considered multiple infection-related hospital admissions occurring within 7 days as the same admission, reflecting possible failed discharge or delayed diagnosis after transfer from the initial medical admissions unit.

Index date was the date of hospital admission. Individuals without any hospitalisation primarily due to infection after age 65 were classified as unexposed and assigned the same index date as their matched exposed counterpart.

### Outcome

The outcome was frailty category or death. Frailty was assessed using the first version of the Electronic Frailty Index (eFI).^26^ Developed and validated in England, the eFI uses routinely collected primary care data to predict all-cause mortality, hospitalisation, and nursing home admission.^26^ It is based on a cumulative deficit framework, with 36 equally weighted deficits.^26^ We defined the polypharmacy deficit as prescriptions of five or more unique drugs on the British National Formulary Chapters1-15.^26^ We identified the other 35 deficits from primary care morbidity records coded using Systematized Nomenclature of Medicine Clinical Terms (SNOMED-CT). We calculated baseline eFI from all deficits recorded through diagnostic codes or prescriptions up to 14 days before the index date (to preclude including acute infection-related change in health status for establishing baseline frailty).

During follow-up, we treated frailty as time-updated and recalculated the eFI whenever a new deficit (based on codelists or prescriptions) was recorded. We classified frailty states using validated cut-points (fit: 0-0.12; mild: >0.12-0.24; moderate: >0.24-0.36; severe: >0.36). For each frailty state, we determined whether participants reached that state during follow-up (yes/no) and recorded the corresponding dates. We obtained death status and date from primary care records. When death and entry into a frailty category occurred on the same date, death was prioritised.

### Covariates

We used directed acyclic graphs to guide selection of potential confounders for the association between severe infection and frailty progression (**Figure S1**). Covariates were measured using information recorded on or before the index date.

We considered the following four broad categories of potential confounders: 1) baseline frailty score; 2) deprivation: individual-level quintiles of IMD, supplemented with practice-level IMD where missing; ^27^ 3) lifestyle factors: harmful alcohol use, and smoking; and 4) primary care infections in the prior 5 years. Ethnicity was included only in a sensitivity analysis because due to missing data for 40% of individuals. We did not adjust for comorbidities as confounders because many comorbid conditions are incorporated within the eFI deficits used to calculate frailty.

We explored potential effect modification by age group, sex, deprivation, care home residency, dementia, diabetes mellitus, as these factors have been linked to infections, frailty, or functional decline.^1,5–12^ We also explored whether transition risk differed by infection pathogen (bacterial, viral, fungal, parasitic) and infection types (sepsis, urinary tract infection [UTI], skin and soft tissue infection [SSTI], meningoencephalitis, lower respiratory tract infection [LRTI], gastroenteritis).

We used previously defined algorithms based on primary care coding to ascertain smoking status^30^ and ethnicity^31^. Harmful alcohol use was based on primary care recording before index date for morbidity codes implying harmful or heavy alcohol use or a prescription for medications used to maintain abstinence. Primary care infection history was based on one relevant SNOMED-CT code.

Care home residency was based on primary care codes suggesting residency in a care or nursing home. We identified antibiotics using the dictionary of medicines and device product codes representing all medications in Section 5.1 of the British National Formulary (excluding antituberculous and antilepromatous agents, and methenamine) because these are specialist mycobacterial treatments or urinary antiseptics rather than routine antibacterial therapy.^32^

### Statistical analysis

We summarised baseline characteristics at index date and overall follow-up time by severe infection status.

We conducted a five-state (fit, mild, moderate, severe, death) Markov multistate model allowing 10 forward transitions from baseline frailty state (fit, mild, moderate, severe) and intermediate states to higher states or the final absorbing state of death (**Figure 1**).^33^ We restricted the model to forward transitions because the frailty measure (eFI) does not capture recovery.

**Figure 1.**
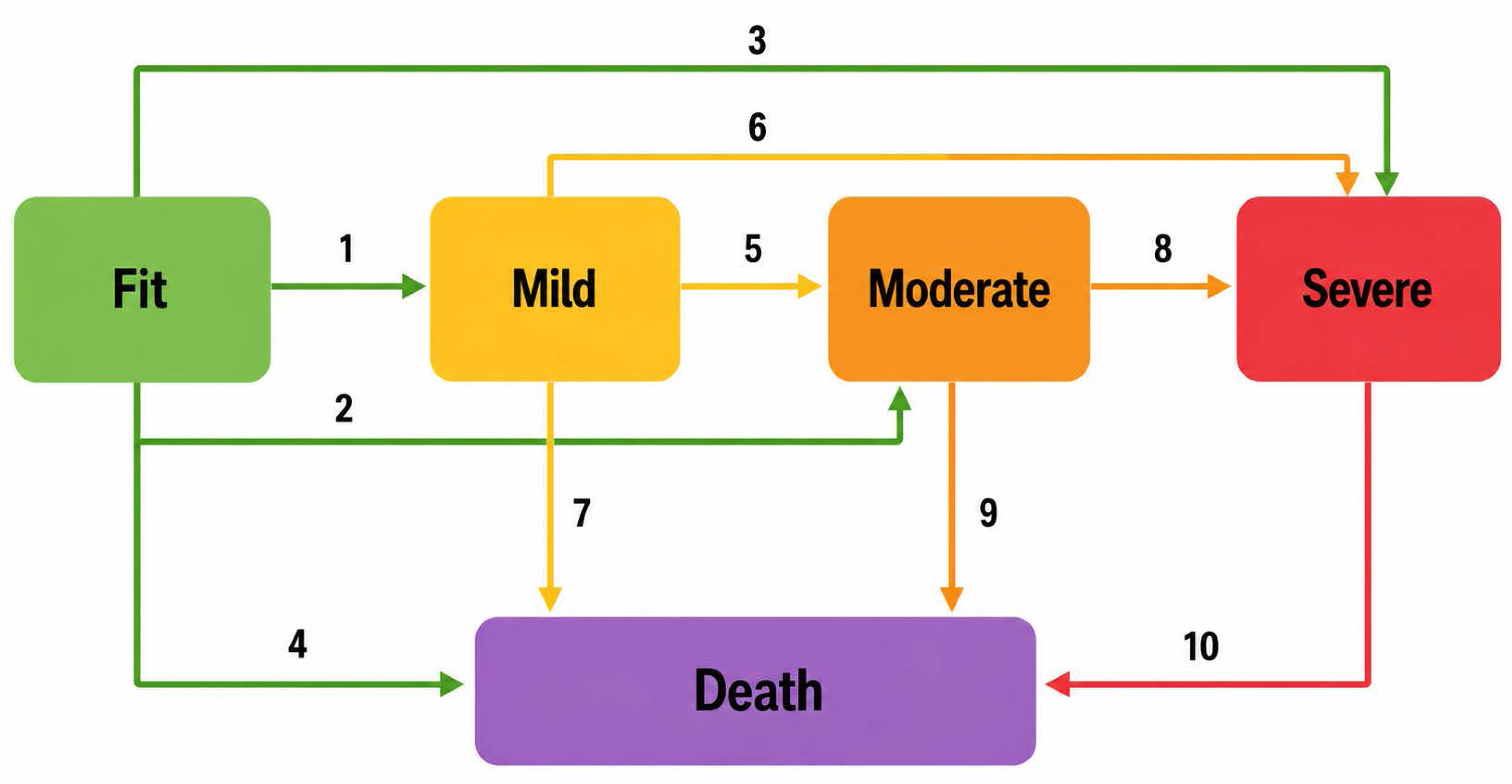
Illustration of a five-state multistate model of 10 transitions to greater frailty or death. At index date individuals are either fit, mild, moderate, or severely frail. Transitions are allowed from the baseline or intermediate states to greater frailty or death. CPRD = Clinical Practice Research Datalink.

For each transition, we estimated cause-specific hazard ratios (HRs) with 95% confidence intervals (CIs) comparing adults with and without severe infection using Cox regression with cluster robust standard errors (by matched-group, hence, implicitly adjusted for matching variables age, sex, and primary care practice). Models were further adjusted for (1) baseline frailty score, and sequentially for other confounders: (2) IMD quintiles, (3) smoking, and harmful alcohol use, and (4) primary care infection history in past five years (fully adjusted model). For transitions originating from the same state, individuals contributed to all possible outgoing transitions, with the same risk sets and survival times, but participants contributed an event only to the transition they experienced (transition=yes) and were censored (transition = no) for transitions not experienced.^33^ From the fully adjusted models, we derived state occupancy probabilities and expected length of stay (ELOS) in each state up to five years of follow-up (based on the average follow-up length) for individuals with and without severe infection.

Under our full Markov assumption, transition hazards depended only on the current state, and hence, the time scale was clock-forward (time since baseline).^33^ We allowed transition hazards to vary over time (i.e., time-inhomogeneous model) and allowed different baseline hazards and covariate effects for each transition.

#### Sensitivity analyses

We evaluated the robustness of our results in three sensitivity analyses based on the fully adjusted Cox regression multistate models. First, to evaluate the impact of residual confounding due to not adjusting for ethnicity we further adjusted for ethnicity. Second, to assess potential exposure misclassification by hospital-acquired infections, we repeated the analysis using a stricter exposure definition that identified infections likely to have been community-acquired within 7 days before the hospital admission as: 1) primary care infection diagnosis; or 2) primary care antibiotic prescription; or 3) both. Third, to assess the influence of temporal changes in recording of eFI deficits and covariates based on the Quality and Outcomes Framework (where some specific primary care morbidity-coded recording were financially incentivised) changes and other similar policy changes over time we restricted to individuals with index dates from 2012 onwards and from 2014 onwards.^34^

#### Secondary analyses

We conducted stratified analyses by levels of potential effect modifiers including age group, sex, quintiles of IMD, care home residency, dementia, diabetes mellitus, infection pathogen, and infection type.

All analyses were performed using R, and the multi-state models were fitted using the mstate package.^35,36^ Analytical code and codelists used to define severe infection (including pathogen and type categories), eFI deficits and covariates are available online (<u>Analytical</u> <u>code</u>, <u>Codelist</u>). The study was approved by the London School of Hygiene & Tropical Medicine Research Ethics Committee (31298) and CPRD’s Independent Scientific Advisory Committee (24_004305).

## Results

We included 1,452,117 adults aged ≥65 years, comprising 243,743 adults with severe infection and 1,208,374 matched comparators without severe infection (**Figure 2**). Age and sex were balanced between groups, reflecting the matching strategy, and ethnicity and deprivation profiles were also largely comparable (**Table 1**). Compared with matched comparators, adults with severe infection had slightly higher recorded levels of harmful alcohol use (2·9% with severe infection vs 1·3% without) and current smoking (20% vs 16%). Adults with severe infection had higher baseline frailty than those without, with fewer being fit (40% with severe infection vs 60% without) and most being mildly frail (36% vs 28%), moderately frail (18% vs 8.9%), and severely frail (6·2% vs 2·2%). The proportion of adults with at least one health care consultation within one year before index date was high in both those with and without severe infection. Adults with severe infection had shorter follow-up (median 2·1 years [IQR 0·5-5·0]) than those without severe infection (4·0 years [1·8-7·2]).

**Figure 2.**
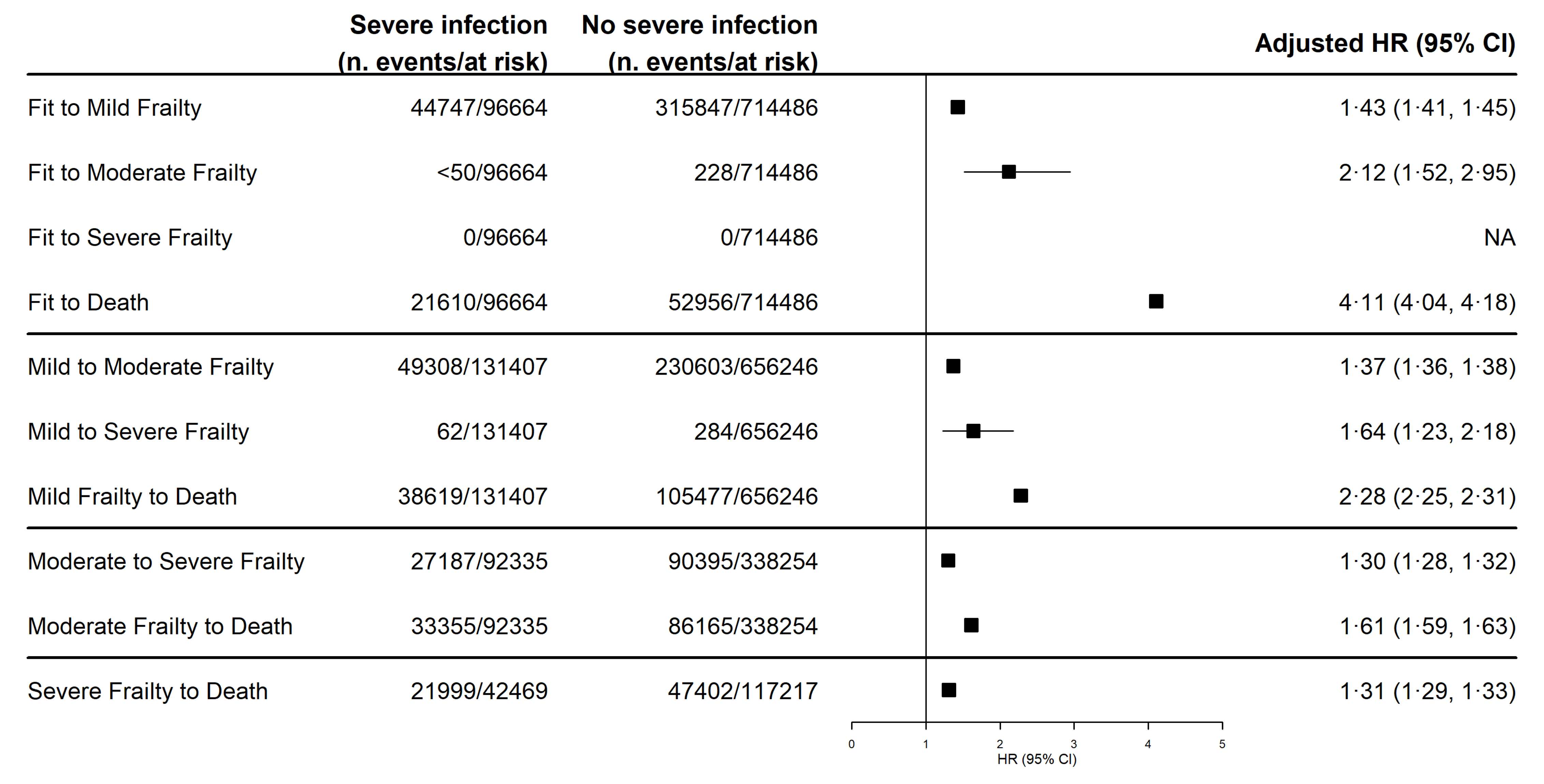
Flow diagram of the study cohort. *Participants were eligible for inclusion from the latest of: study start (01-04-2006); 65th birthday; or one year after registration with practice. LRTI=Lower Respiratory Tract Infection, SSTI=Skin and Soft Tissue Infection, UTI=Urinary Tract Infection.

**Table 1.** Characteristics of adults at index date and during follow-up.

|  | **No Severe infection,**<br>**n = 1,208,374** | **Severe infection,**<br>**n = 243,743** |
| --- | --- | --- |
| **Age, years** |  |  |
| Median (IQR) | 76 (70, 84) | 77 (70, 84) |
| 65-70 | 337,556 (28%) | 67,342 (28%) |
| 71-80 | 444,179 (37%) | 88,602 (36%) |
| 81-90 | 350,077 (29%) | 70,319 (29%) |
| >90 | 76,562 (6·3%) | 17,480 (7·2%) |
| **Sex** |  |  |
| Female | 665,097 (55%) | 133,951 (55%) |
| Male | 543,277 (45%) | 109,792 (45%) |
| **Ethnicity** |  |  |
| White | 650,450 (54%) | 135,268 (55%) |
| South Asian | 28,303 (2·3%) | 5,413 (2·2%) |
| Black | 18,162 (1·5%) | 3,050 (1·3%) |
| Other | 5,610 (0·5%) | 715 (0·3%) |
| Mixed | 3,792 (0·3%) | 640 (0·3%) |
| Missing | 502,057 (42%) | 98,657 (40%) |
| **Index of Multiple Deprivation (quintiles)** |  |  |
| 1, least deprived | 272,382 (23%) | 51,274 (21%) |
| 2 | 261,959 (22%) | 52,414 (22%) |
| 3 | 245,302 (20%) | 48,697 (20%) |
| 4 | 226,489 (19%) | 47,097 (19%) |
| 5, most deprived | 202,242 (17%) | 44,261 (18%) |
| **Harmful alcohol use** | 16,137 (1·3%) | 6,950 (2·9%) |
| **Smoking status** |  |  |
| Current | 195,290 (16%) | 48,933 (20%) |
| Former | 356,345 (29%) | 83,067 (34%) |
| Never | 637,211 (53%) | 109,529 (45%) |
| Missing | 19,528 (1·6%) | 2,214 (0·9%) |
| **Care home residency** | 27,230 (2·3%) | 4,731 (1·9%) |
| **Primary care recorded infection**<br>**(5 years lookback)** | 546,750 (45%) | 173,887 (71%) |
| **Primary care recorded infection**<br>**(7 days lookback)** | 7,866 (0·7%) | 58,019 (24%) |
| **Primary care antibiotic prescription (7 days**<br>**lookback)** | 22,520 (1·9%) | 40,366 (17%) |
| **Dementia** | 71,319 (5·9%) | 20,864 (8·6%) |
| **Diabetes mellitus** | 212,382 (18%) | 62,090 (25%) |
| **Electronic frailty index** |  |  |
| Mean (SD) | 0·12 (0·09) | 0·17 (0·11) |

| **Table 1. Characteristics of adults at index date and during follow-up** |  |  |
| --- | --- | --- |
|  | **No Severe infection,**<br>**n = 1,208,374** | **Severe infection,**<br>**n = 243,743** |
| Fit: 0-0.12 | 731,026 (60%) | 98,386 (40%) |
| Mild: >0.12-0.24 | 342,833 (28%) | 87,065 (36%) |
| Moderate: >0.24-0.36 | 107,913 (8.9%) | 43,061 (18%) |
| Severe: >0.36 | 26,602 (2.2%) | 15,231 (6.2%) |
| At least one healthcare consultation within 12 months before index date | 1,174,951 (97%) | 241,762 (99%) |
| Data are n (%), mean (SD), or median (IQR).<br>All percentages were calculated with the total number in the respective column headers as denominators.<br>Percentages may not add up to 100 due to rounding.<br>* Censoring was due to death, study end, end of participant's registration with practice, or end of data collection from practice.<br>IQR = Interquartile Range, SD = Standard Deviation. |  |  |

In the multi-state models, transition hazard ratios comparing those with and without severe infection did not change after sequential adjustment for confounders (**Figure S2**). After full adjustment for age and sex (based on matching), baseline frailty score, deprivation, smoking, harmful alcohol use, and prior infection, severe infection was associated with higher hazards of transitioning to advanced frailty states or death across all transitions (**Figure 3**). From the fit state, hazards ratios (95% CI) were (to mild frailty: 1·43, 1·41, 1·45; to moderate frailty: 2·12, 1·52, 2·95, and to death: 4·11, 4·04, 4·18) with no transitions to severe frailty. Similar patterns were observed for transitions from mild frailty (to moderate frailty: 1·37, 1·36, 1·38, to severe frailty: 1·64, 1·23, 2·18, to death: 2·28, 2·25, 2·31), from moderate frailty (to severe frailty: 1·30, 1·28, 1·32, and to death: 1·61, 1·59, 1·63), and from severe frailty to death (1·31, 1·29, 1·33). Among transitions from the same frailty state, hazard ratios increased progressively with advancing destination frailty state.

**Figure 3.**
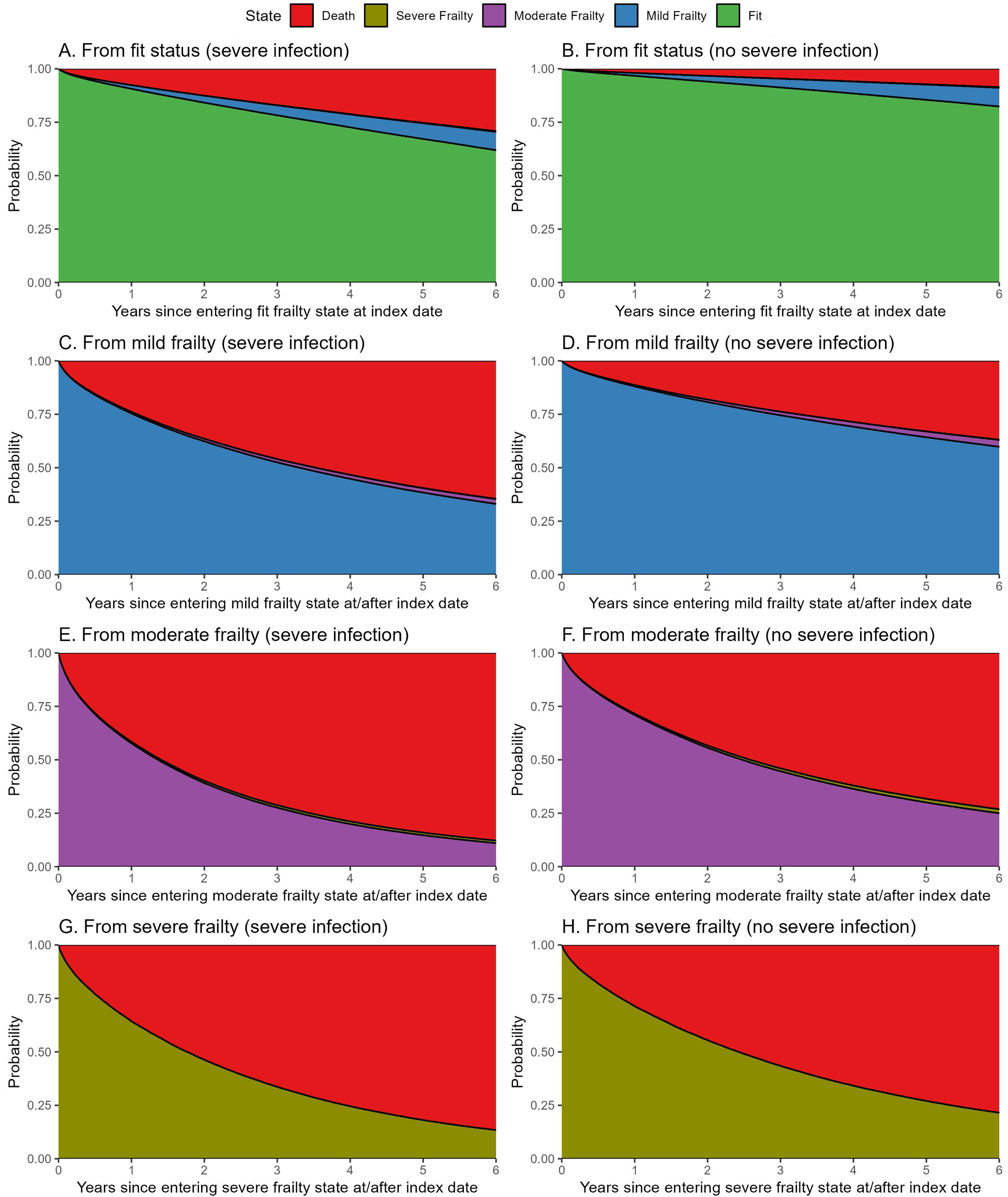
Forest plots of transition hazard ratios from the multistate model comparing adults with and without severe infection. Estimates are from separate Cox regression models for each transition adjusted for age, sex, deprivation, smoking, harmful alcohol use, and prior infection. CI = Confidence Interval, HR = Hazard Ratio.

Hazard ratios for transitions to death were highest from the fit state and decreased with increasing frailty severity. But cumulative incidence proportions for death were lowest among fit individuals and increased progressively across frailty states. This reflects higher underlying mortality risk among individuals with greater frailty, despite smaller relative effect estimates by severe infection exposure.

The probability of remaining in each frailty state (i.e., not transitioning to greater frailty or death) declined more rapidly over time among adults with severe infection than matched counterparts without severe infection (**Figure 4**). At five years, adults with severe infection had lower probabilities of remaining in each frailty state, fit (67·2% with severe infection vs 85·4% without), mild frailty (38·4% vs 64·3%), moderate frailty (14·7% vs 30·0%), and severe frailty (18·1% vs 27·0%) and hence had higher probabilities of transitioning to advanced frailty states or death than those without severe infection (**Table S1**). Similarly, adults with severe infection had shorter expected length of stay in each frailty state at five years, fit (4·1 years with severe infection vs 4·6 years without), mild frailty (3·0 vs 3·9) moderate frailty (2·0 years vs 2·7), and severe frailty (2·2 vs 2·6) (**Table S2**).

**Figure 4.**
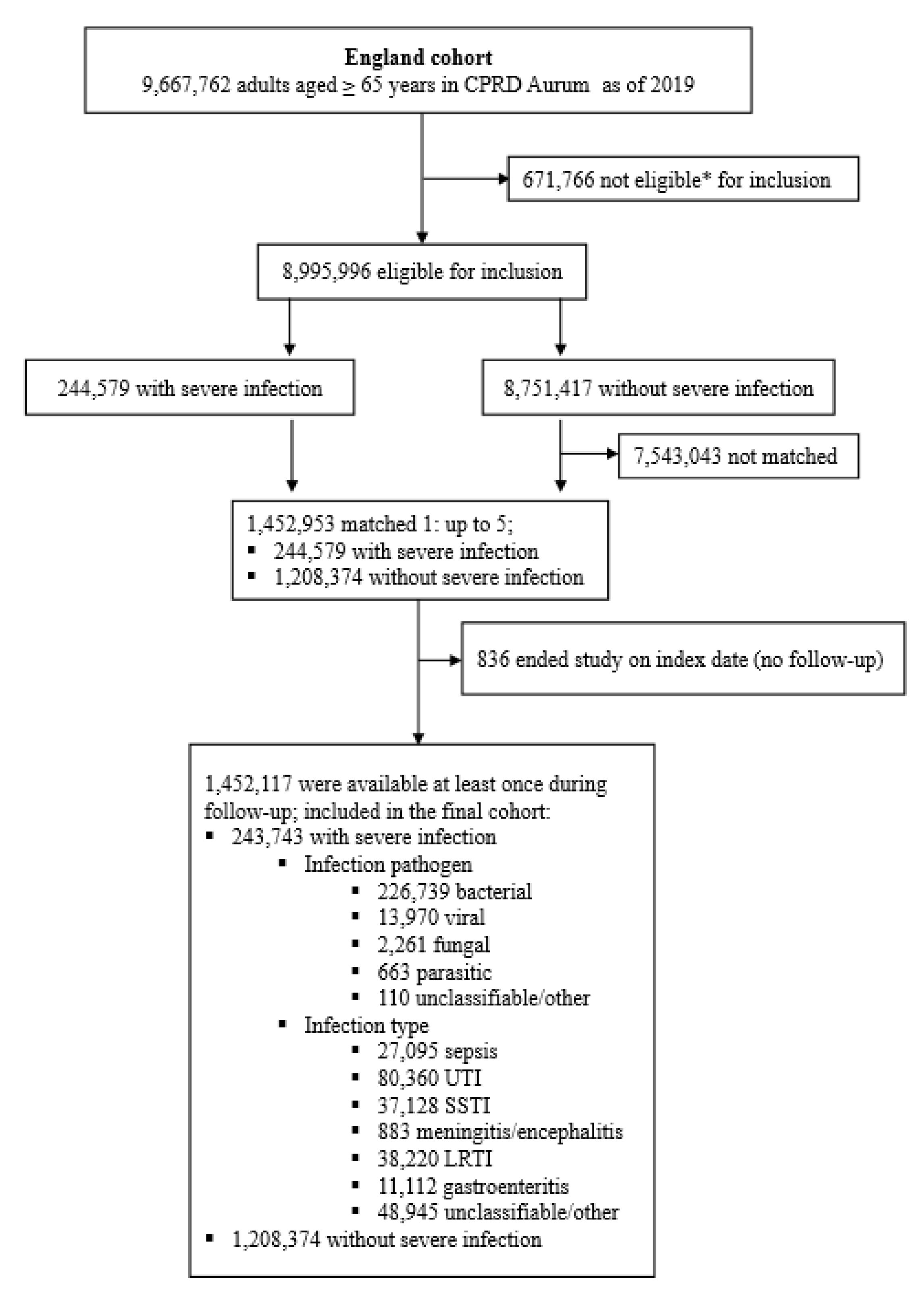
Stacked state occupancy probabilities over time in participants with and without severe infection. Estimates are from separate Cox regression models for each transition adjusted for age, sex, deprivation, smoking, harmful alcohol use, and prior infection. The shaded area indicates the probability of being in a state.

### Sensitivity analyses

Results of sensitivity analyses were broadly consistent with those of the main analyses, including further adjustment for ethnicity, and restriction to Quality and Outcomes Framework periods after 2012, and after 2014 (**Figures S3-S5)**. However, hazard ratios (95% CI) for transitions from fit to moderate frailty were higher when infections appeared to be community-acquired 7 days before index date, as indicated by a primary care diagnosis (9·31, 1·10-79·02), or a primary care diagnosis plus antibiotic prescription (7·46, 1·84-30·22) (**Figure S4**).

### Secondary analyses

Hazard ratios (95% CI) for transitions from fit to death were highest at younger ages and decreased with increasing age (65-70 years: 7·06, 6·75-7·37; 71-80 years: 6·45, 6·19-6·73; 81-90 years: 4·48, 4·26-4·72, and >90 years: 2·63, 2·48-2·78) with similar patterns observed from mild frailty to death (**Figure S6**). Hazard ratios for all other transitions were similar by age group.

Hazard ratios were lower among adults residing in care homes (**Figure S9**). Apart from mild to severe frailty, all transition hazard ratios were lower among adults among adults with dementia, fit to (mild[1·22, 1·15, 1·31], moderate[0·57, 0·08, 3·94], death[1·83, 1·72, 1·95]), than adults without dementia, fit to (mild[1·43, 1·42, 1·45], moderate[2·21, 1·58, 3·10], death[4·29, 4·22, 4·37]) (**Figure S10**).

Transition hazard ratios to higher frailty varied by type of infection exposure, highest with LRTI, fit to (mild[1·91, 1·84, 1·98], moderate[3·86, 1·73, 8·59], death[7·15, 6·82, 7·49, followed by sepsis, fit to (mild[1·76, 1·67, 1·85], moderate[1·24, 0·16, 9·65], death[14·73, 14·07, 15·43]) and then followed by urinary tract infections, meningitis/encephalitis, gastroenteritis, and skin and soft tissue infections (**Figure S13**). Transitions from frailty states to death were highest with sepsis, followed by LRTI, meningitis/encephalitis, gastroenteritis, UTI, SSTI.

There was limited evidence of effect modification by, sex, deprivation, diabetes, or infection pathogen (**Figures S6-S12**).

## Discussion

From a large-scale national electronic health records database, we found that severe infection, defined as hospitalisation primarily due to infection after age 65 years, increased the risk of progression from baseline and intermediate frailty states to more advanced frailty states or death. Among transitions starting from the same frailty state, the risk increased progressively to more advanced destination states, including death. When considering only transitions to death, the risk was highest from the fit state and declined progressively with each successive starting frailty state. At five years, severe infection was associated with lower probabilities of remaining in each frailty state and lower expected time spent in each state. The risk of progression to greater frailty or death varied by infection type, with the strongest associations observed for LRTI, followed by sepsis, UTI, meningitis/encephalitis, gastroenteritis, and skin and soft tissue infections. Transitions to death were highest with sepsis, followed by LRTI, meningitis/encephalitis, gastroenteritis, UTI, and SSTI. Our findings support the hypothesis that severe infections in old age are associated with more rapid progression to greater frailty or death.

We found only a handful of longitudinal studies investigating infection-related frailty progression. Their findings were broadly consistent with those in our study, although they often focussed on specific infections and rarely accounted for competing risk of death. These studies reported higher risk of transition from a fit or pre-frail state to frailty after SARS-CoV-2,^24^ HIV,^37^ and a broad range of 20 infections,^14^ including influenza, tuberculosis, urinary tract infections, sexually transmitted infections, and lower and upper respiratory tract infections. Studies examining infection history using antibody concentration or seropositivity, that reported increased risks of transition from fit/pre-frail states to frailty, are also consistent with our study.^10,38^

Unlike previous studies, our study included a broad range of recorded infections and used a five-state multistate model to examine all transitions to greater frailty while accounting for competing risk of death. We used a large, routinely collected dataset from a national health care system that is broadly representative of the source population. We assessed whether transition rates varied by pathogen and infection type. We also adjusted for a wide range of potential confounders, including baseline frailty, and our findings were robust across sensitivity analyses.

Our definition of severe infection was based on hospital admissions in which infection was recorded as the primary diagnosis. This may have led to exposure misclassification if some individuals classified as comparators had severe infections recorded in other diagnostic positions, potentially attenuating differences in frailty progression between groups. In addition, people with community-managed infections or infections that did not result in hospital admission would have been classified as unexposed. However, infections managed outside hospital were likely to be less severe. Thus, any resulting misclassification would therefore probably have biased our estimates towards the null. Further, some infections may have been hospital-acquired and therefore misclassified as severe infections. Reassuringly, findings were consistent in sensitivity analyses restricted to adults with evidence of community-acquired infection and their matched comparators.

Because our frailty measure was cumulative and could only increase over time, we were unable to investigate frailty recovery, defined as movement from more advanced to less severe frailty states.^26^ Although modelling frailty recovery could provide additional insight into longer-term post-infection trajectories, our primary focus was on the adverse public health impact of infections on frailty progression.

Also, participants with severe infection may have had more healthcare contacts over time, increasing the likelihood that frailty deficits were recorded. This differential ascertainment of frailty status between exposed and unexposed groups may have led to overestimation of the association between severe infection and frailty progression. However, most adults in both groups had at least one consultation within 12 months before the index date, indicating potentially similar healthcare contacts over time among adults with and without severe infection.

The unexpected finding of slower frailty progression after infection among individuals with dementia may also reflect differential frailty ascertainment. People with dementia may be less likely to report symptoms, seek healthcare, or attend scheduled appointments, reducing opportunities for new deficits to be recorded. In addition, clinicians may have less incentive to document or investigate accumulating frailty deficits in individuals with established dementia, where the clinical or social care implications of further deficit recording may be limited. Future studies using alternative frailty measures that are less reliant on healthcare-seeking behaviour may help to further clarify this relationship.

We cannot exclude residual confounding. Some relevant factors are not captured consistently in routinely collected health records, including social engagement, carer support, and access to occupational or physical therapy. These factors may influence both the risk of severe infection and subsequent frailty progression but could not be robustly accounted for in our analyses.

Nevertheless, our findings suggest that older adults may progress more rapidly to advanced frailty states after severe infection than comparable adults without severe infection. The rate of progression also appeared to vary by infection type, with the strongest associations observed after LRTI, followed by sepsis, UTI, meningitis/encephalitis, gastroenteritis, and skin and soft tissue infections. These infection groups may therefore represent priority targets for prevention and post-infection interventions aimed at mitigating frailty deterioration. Infection prevention, including vaccination, alongside early detection and prompt management, could help reduce frailty progression in older age. Frailty progression is multifaceted, and further research is needed to understand its broader life-course determinants and inform more holistic prevention strategies.

## Data Availability

We do not have the rights to publicly share the data. Data are available on request from CPRD in the UK.

https://gateway.cprd.com/

## Contributors

KA, KEM, CTR, and CWG conceived the study and contributed to its design. KA, KEM, and CWG prepared the statistical analysis plan. KA undertook data extraction and verification. KA, KEM, GRGL, EB, RK, CTR, and CWG had data access through their affiliation with the London School of Hygiene & Tropical Medicine. KA was responsible for data management, cleaning, and analyses. KEM, and CWG provided oversight of the statistical analyses. KA, KEM, and CWG prepared the first draft of the manuscript. All authors contributed to interpretation of the findings and critically reviewed and approved the final manuscript. All authors accept responsibility for the decision to submit the manuscript for publication.

## Acknowledgements

This study used data from the Clinical Practice Research Datalink, obtained under licence from the UK Medicines and Healthcare products Regulatory Agency. Data are contributed by patients and collected by the National Health Service as part of routine care and support. This work was funded through a Wellcome Career Development Award to CWG (225868/Z/22/Z).

