## Supplementary material for "A multistate model of frailty progression after severe infections in adults ≥65 years in England: a matched-cohort study"

**Supplementary Appendix 1: Supplementary Tables**

| **Table S1. State occupancy probabilities at Year 5 follow-up** | | | | | | |
| --- | --- | --- | --- | --- | --- | --- |
|  | | **To** | | | | |
|  | **From** | **Fit**  **% (95% CI)** | **Mild**  **% (95% CI)** | **Moderate**  **% (95% CI)** | **Severe**  **% (95% CI)** | **Death**  **% (95% CI)** |
| **Severe infection** | **Fit** | 67·2  (66·6, 67·8) | 7·3  (7·1, 7·5) | 0·2  (0·2, 0·2) | 0 | 25·3  (24·7, 25·9) |
|  | **Mild** | ·· | 38·4  (37·6, 39·2) | 2·1  (2·1, 2·1) | 0·1  (0·1, 0·1) | 59·5  (58·5, 60·5) |
|  | **Moderate** | ·· | ·· | 14·7  (13·9, 15·5) | 1·2  (1·2, 1·2) | 84·0  (83, 85) |
|  | **Severe** | ·· | ·· | ·· | 18·1  (16·7, 19·5) | 81·9  (80·5, 83·3) |
| **No severe infection** | **Fit** | 85·4  (85·2, 85·6) | 7·2  (7, 7·4) | 0·2  (0·2, 0·2) | ·· | 7·2 (7, 7·4) |
|  | **Mild** | ·· | 64·3  (63·7, 64·9) | 2·7  (2·7, 2·7) | 0·1  (0·1, 0·1) | 33·0  (32·4, 33·6) |
|  | **Moderate** | ·· | ·· | 30·0  (29, 31) | 1·7  (1·7, 1·7) | 68·2  (67·2, 69·2) |
|  | **Severe** | ·· | ·· | ·· | 27·0  (25·6, 28·4) | 73·0  (71·6, 74·4) |

| **Table S2. Expected years of stay in each state at Year 5 follow-up** | | | | | | |
| --- | --- | --- | --- | --- | --- | --- |
|  | | **To** | | | | |
|  | **From** | **Fit** | **Mild** | **Moderate** | **Severe** | **Death** |
| **Severe infection** | **Fit** | 4·1 | 0·2 | ·· | 0 | 0·7 |
|  | **Mild** | ·· | 3·0 | 0·1 | ·· | 1·9 |
|  | **Moderate** | ·· | ·· | 2·0 | 0·1 | 3 |
|  | **Severe** | ·· | ·· | ·· | 2·2 | 2·8 |
| **No severe infection** | **Fit** | 4·6 | 0·2 | ·· | ·· | 0·2 |
|  | **Mild** | ·· | 3·9 | 0·1 | ·· | 1 |
|  | **Moderate** | ·· | ·· | 2·7 | 0·1 | 2·3 |
|  | **Severe** | ·· | ·· | ·· | 2·6 | 2·4 |

**Supplementary Appendix 2: Supplementary Figures**

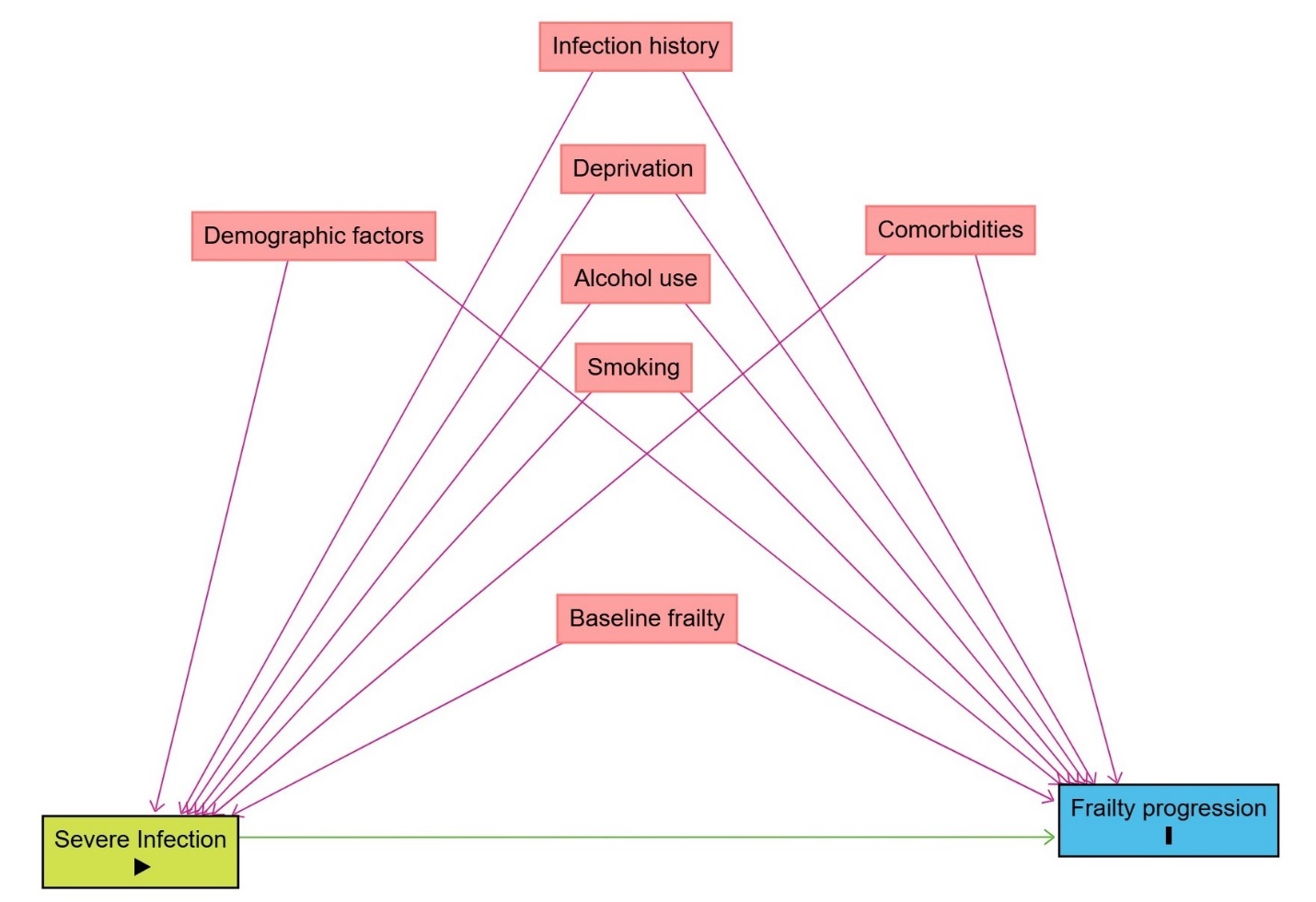

**Figure S1.** Directed acyclic graphic depicting potential confounders of the association between severe infections and frailty progression.

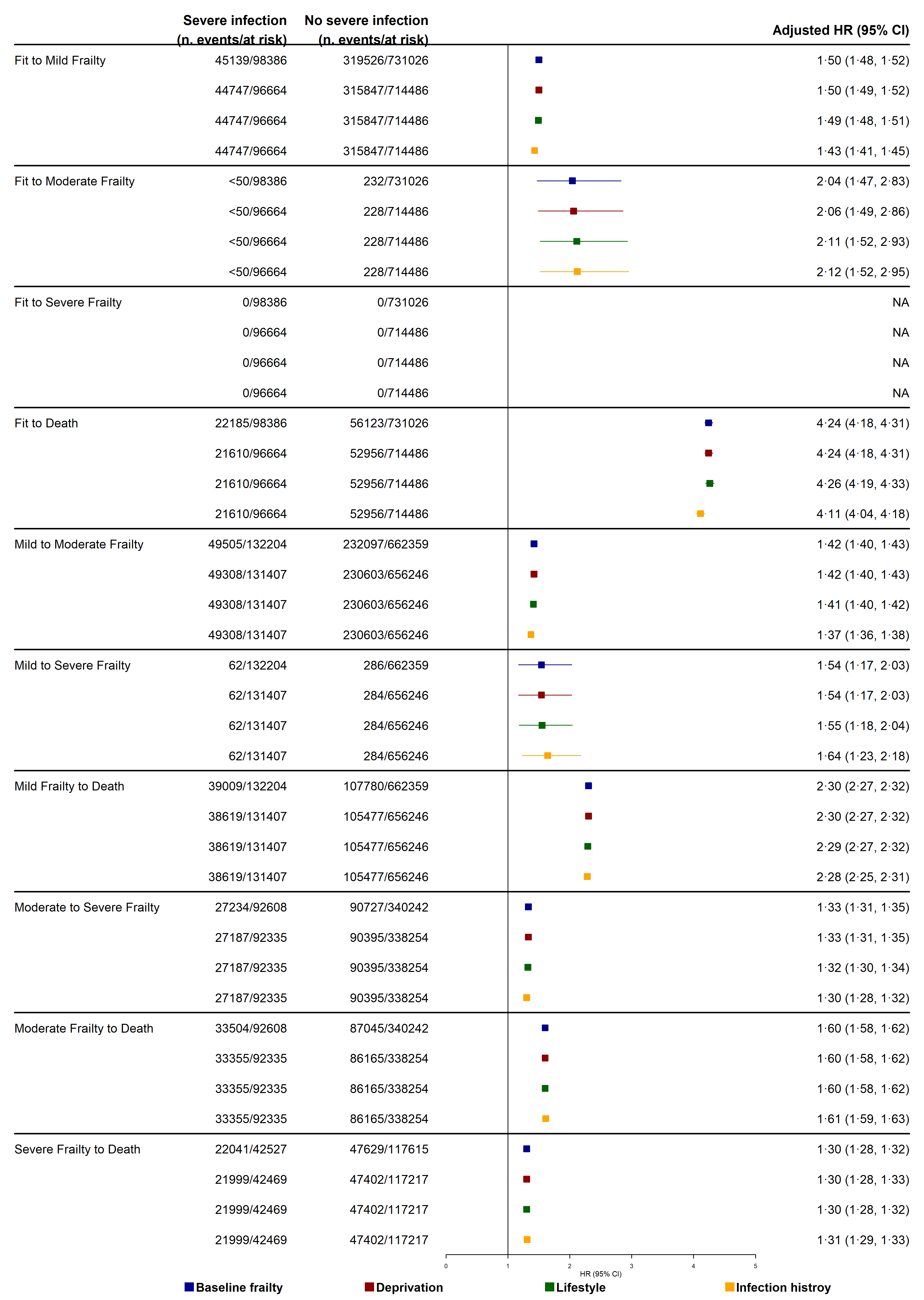

**Figure S2** Forest plots of transition hazard ratios from sequentially confounder-adjusted multistate models comparing adults with and without severe infection. Confounder adjustment sets are Baseline frailty implicitly adjusted for matching factors; Deprivation: Index of Multiple Deprivation; Lifestyle: smoking and harmful alcohol use; and Infection history: primary care recorded infection five years before index date.

CI = Confidence Interval, HR = Hazard Ratio.

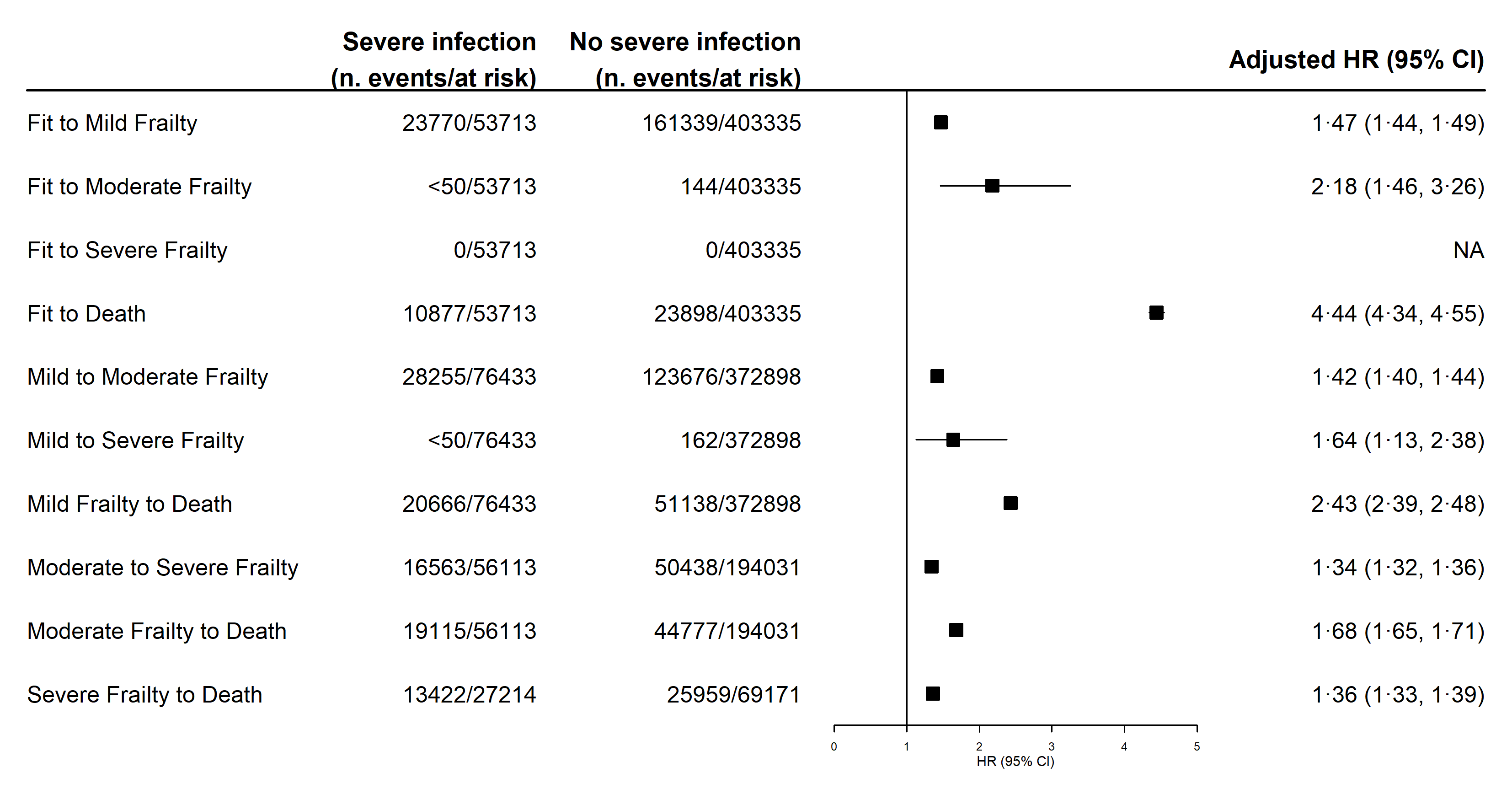

**Figure S3** Forest plots of transition hazard ratios from the multistate model further adjusted for ethnicity comparing adults with and without severe infection.

CI = Confidence Interval, HR = Hazard Ratio.

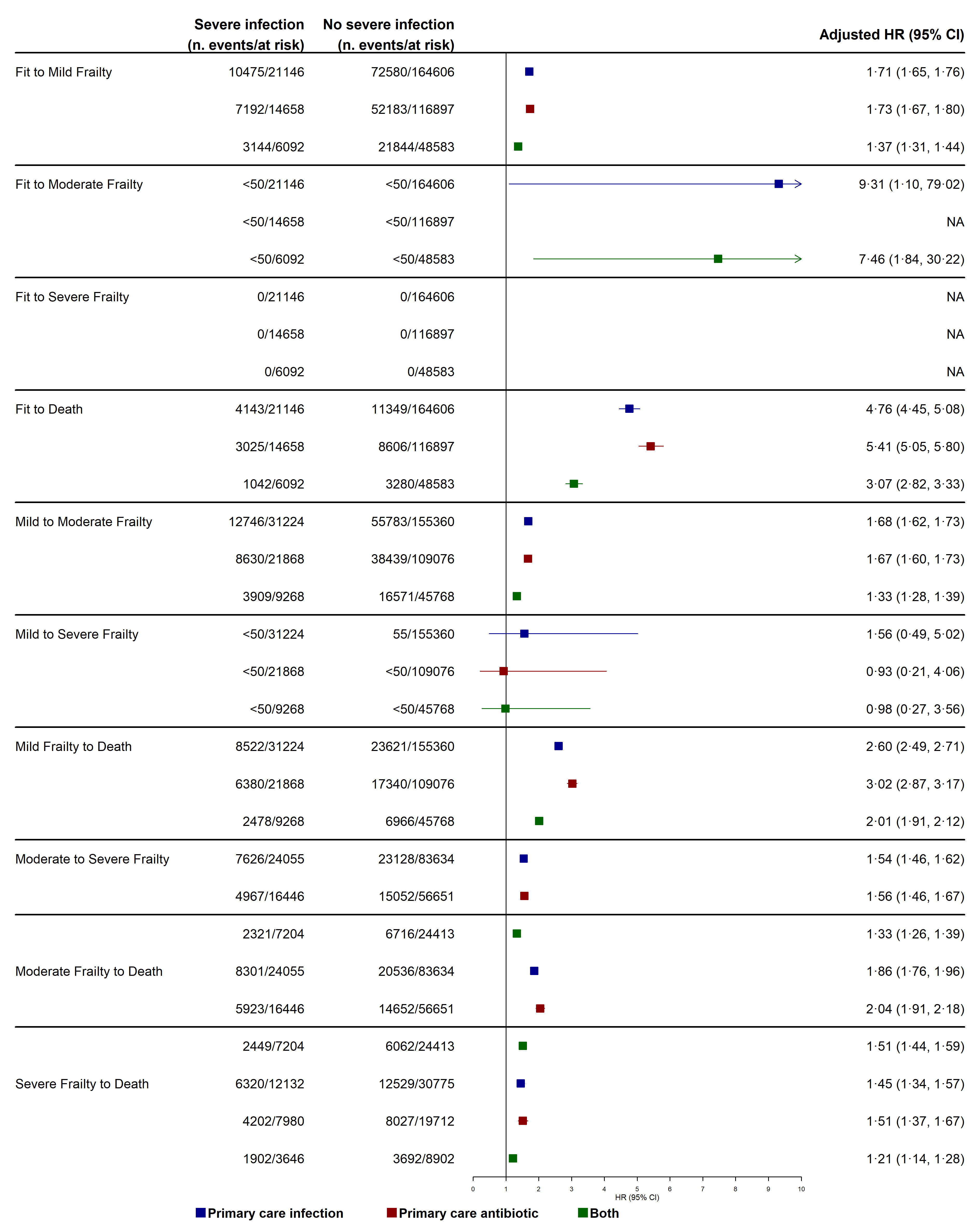

**Figure S4** Forest plots of transition hazard ratios from the multistate model comparing adults with and without severe infection. For each transition, model was restricted to exposed participants with records implying community acquired infection exposure and their matched counterparts defined as: Primary care infection 7 days before index date, or Primary care antibiotic prescribed 7 days before index date, or Both.

CI = Confidence Interval, HR = Hazard Ratio.

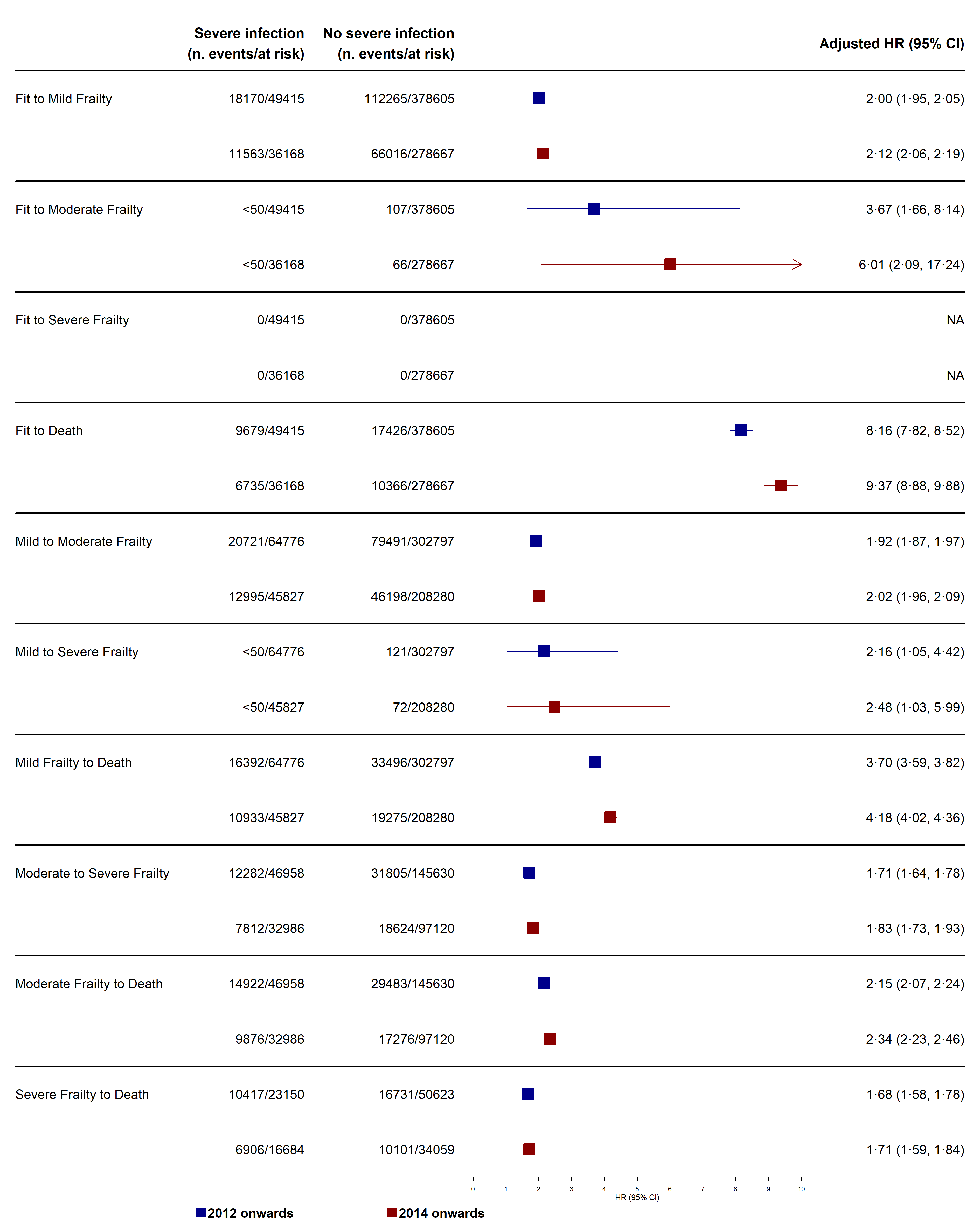

**Figure S5** Forest plots of transition hazard ratios from the multistate model comparing adults with and without severe infection restricted to Quality Outcomes Framework periods: Index date from 2012 onwards, and Index date from 2014 onwards.

CI = Confidence Interval, HR = Hazard Ratio.

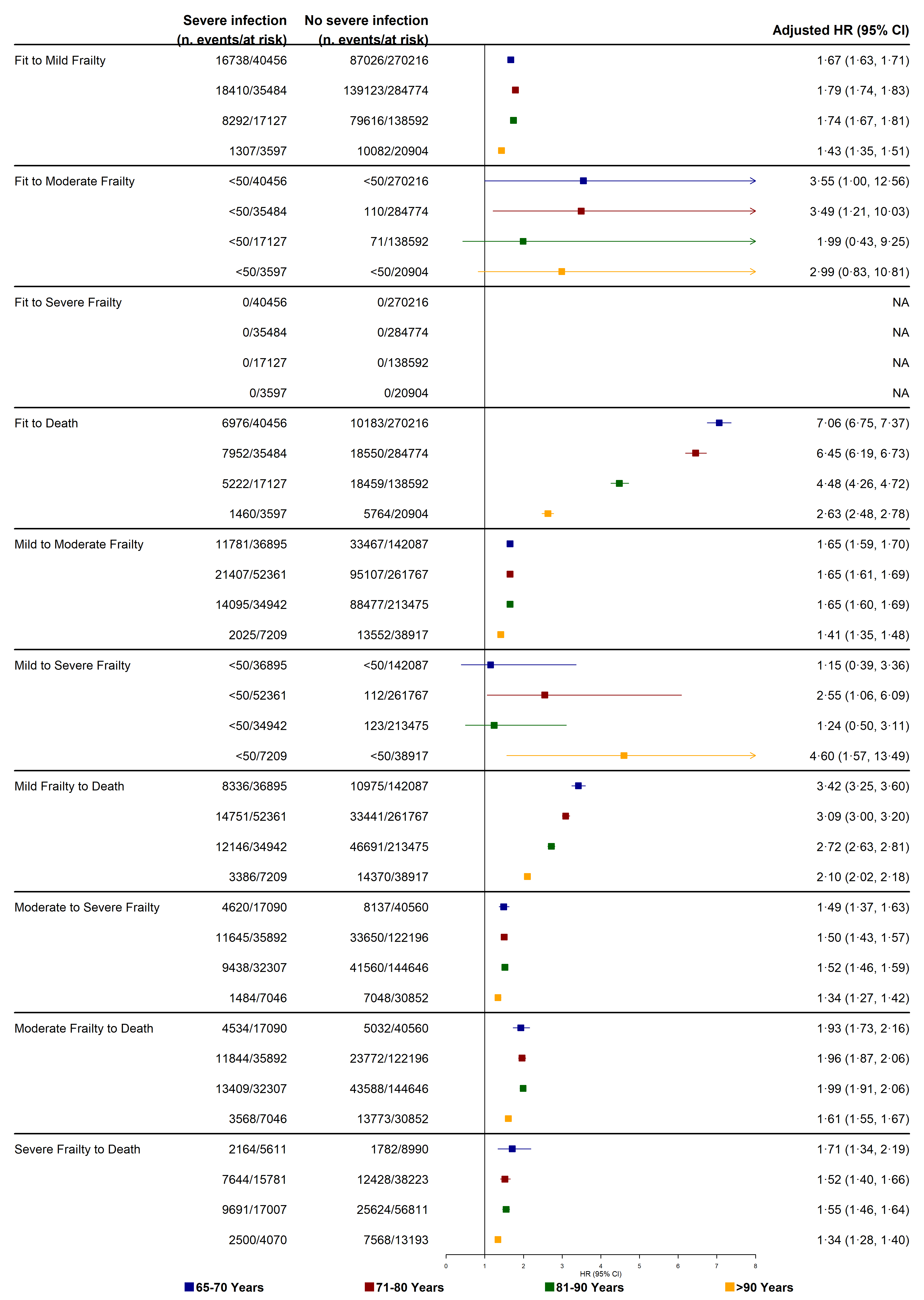

**Figure S6** Effect modification by age group. Forest plots of transition hazard ratios from the multistate model comparing adults with and without severe infection stratified by age groups in years: 65-70, 71-80, 81-90, >90.

CI = Confidence Interval, HR = Hazard Ratio.

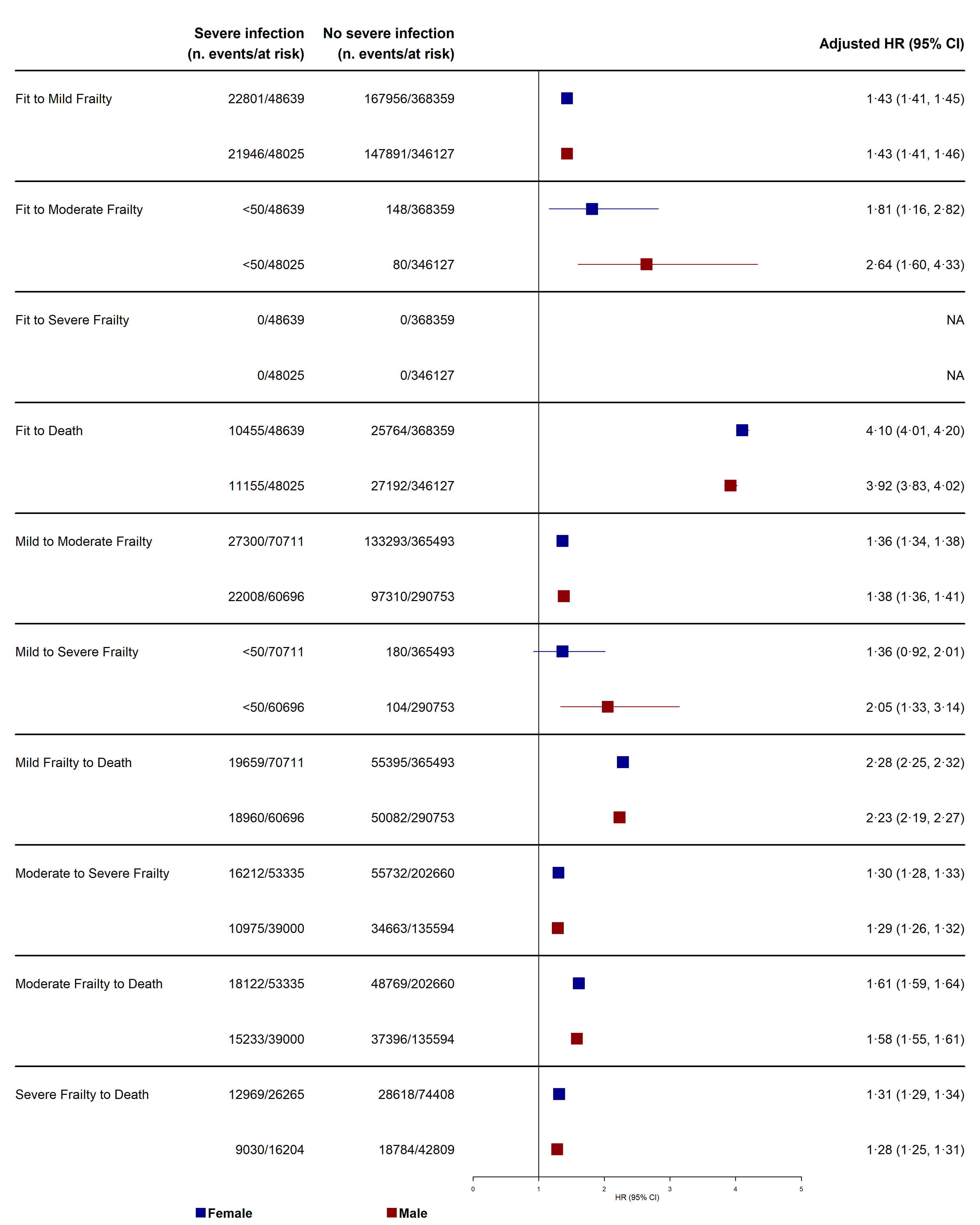

**Figure S7** Effect modification sex. Forest plots of transition hazard ratios from the multistate model comparing adults with and without severe infection stratified by Female, and Male sex.

CI = Confidence Interval, HR = Hazard Ratio.

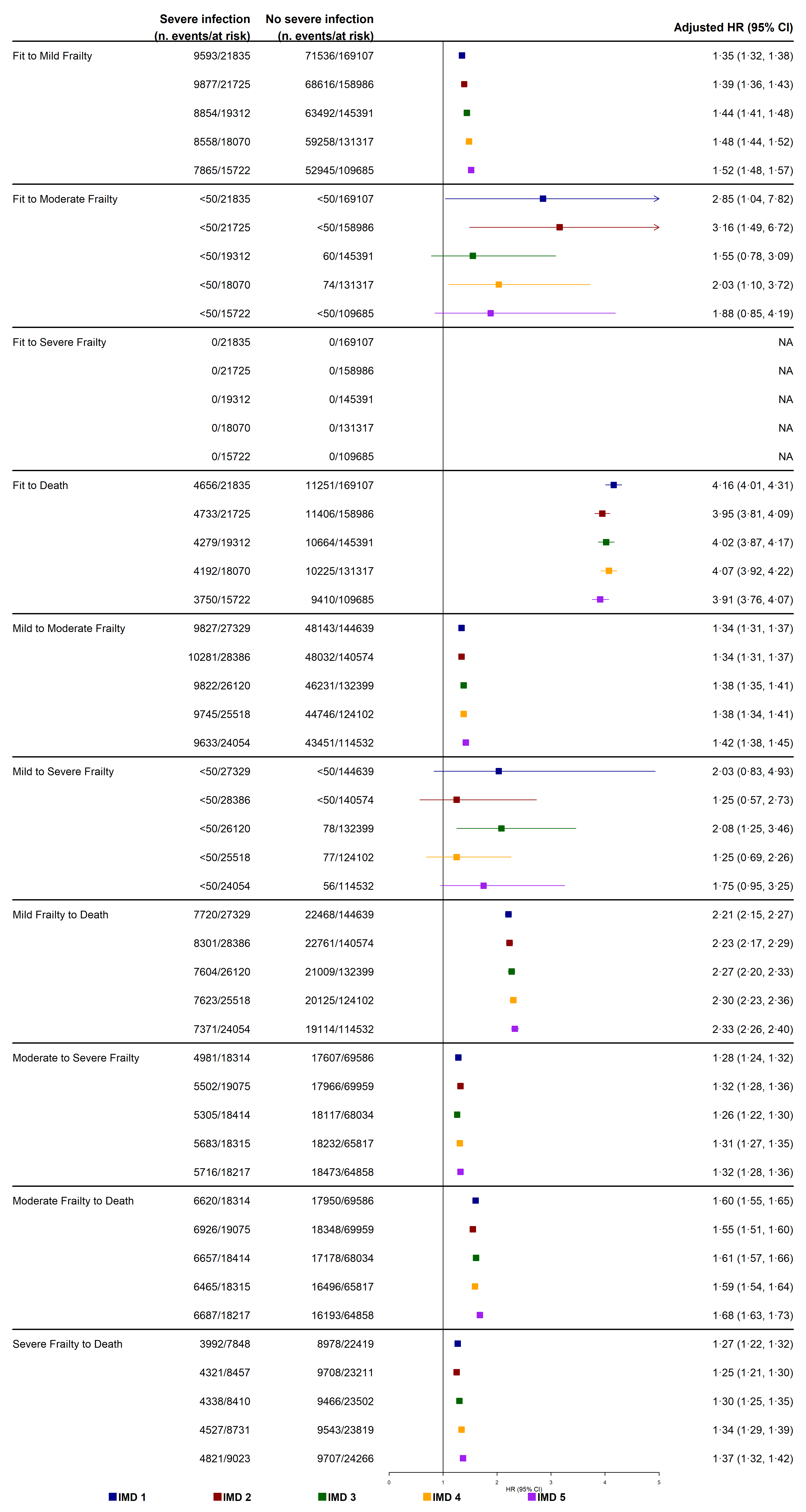

**Figure S8** Effect modification by deprivation. Forest plots of transition hazard ratios from the multistate model comparing adults with and without severe infection stratified by IMD quintiles: 1 (least deprived), 2, 3, 4, and 5 (most deprived).

CI = Confidence Interval, HR = Hazard Ratio, IMD= Index of Multiple Deprivation.

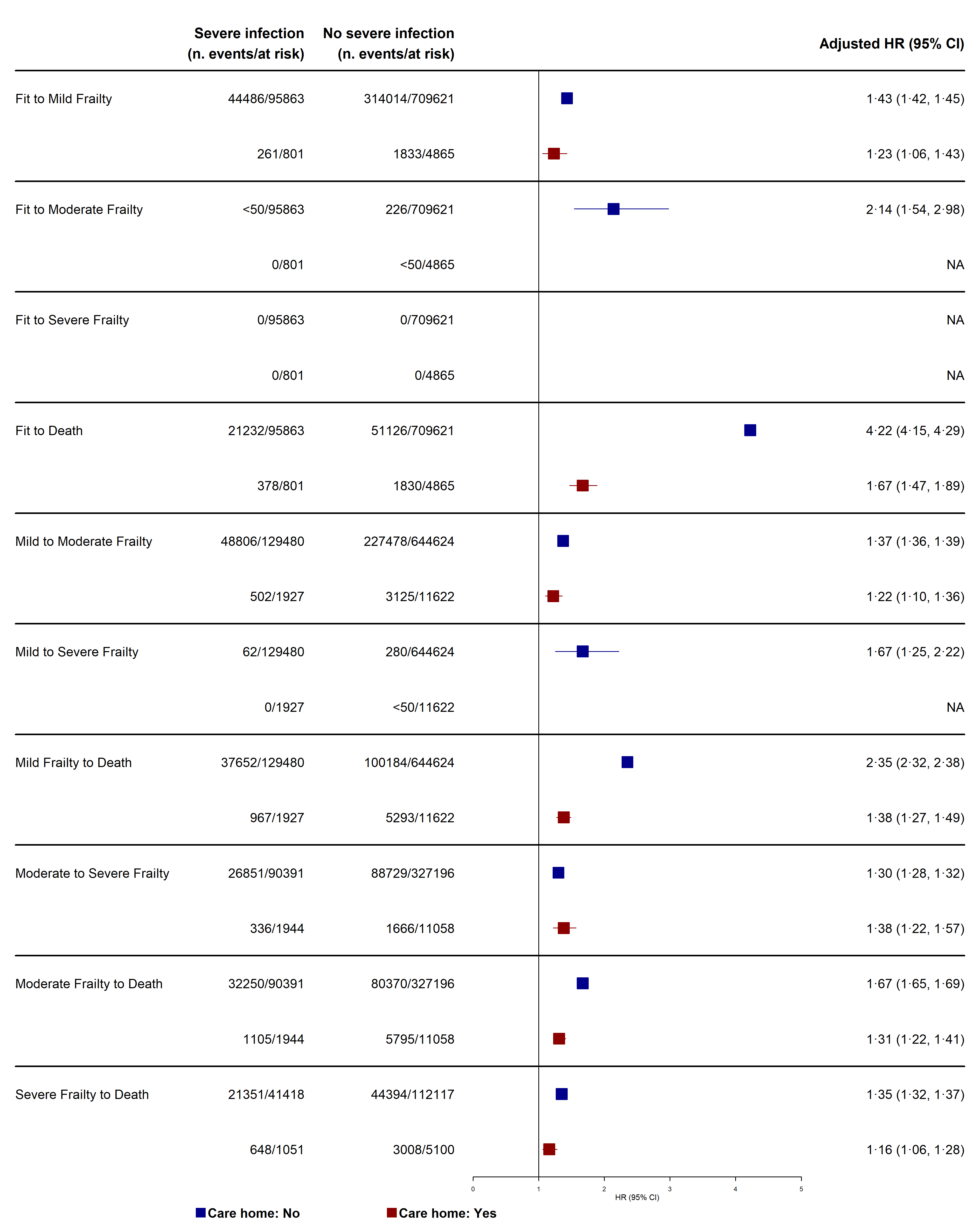

**Figure S9** Effect modification by care home residency. Forest plots of transition hazard ratios from the multistate model comparing adults with and without severe infection stratified by care home residency status.

CI = Confidence Interval, HR = Hazard Ratio.

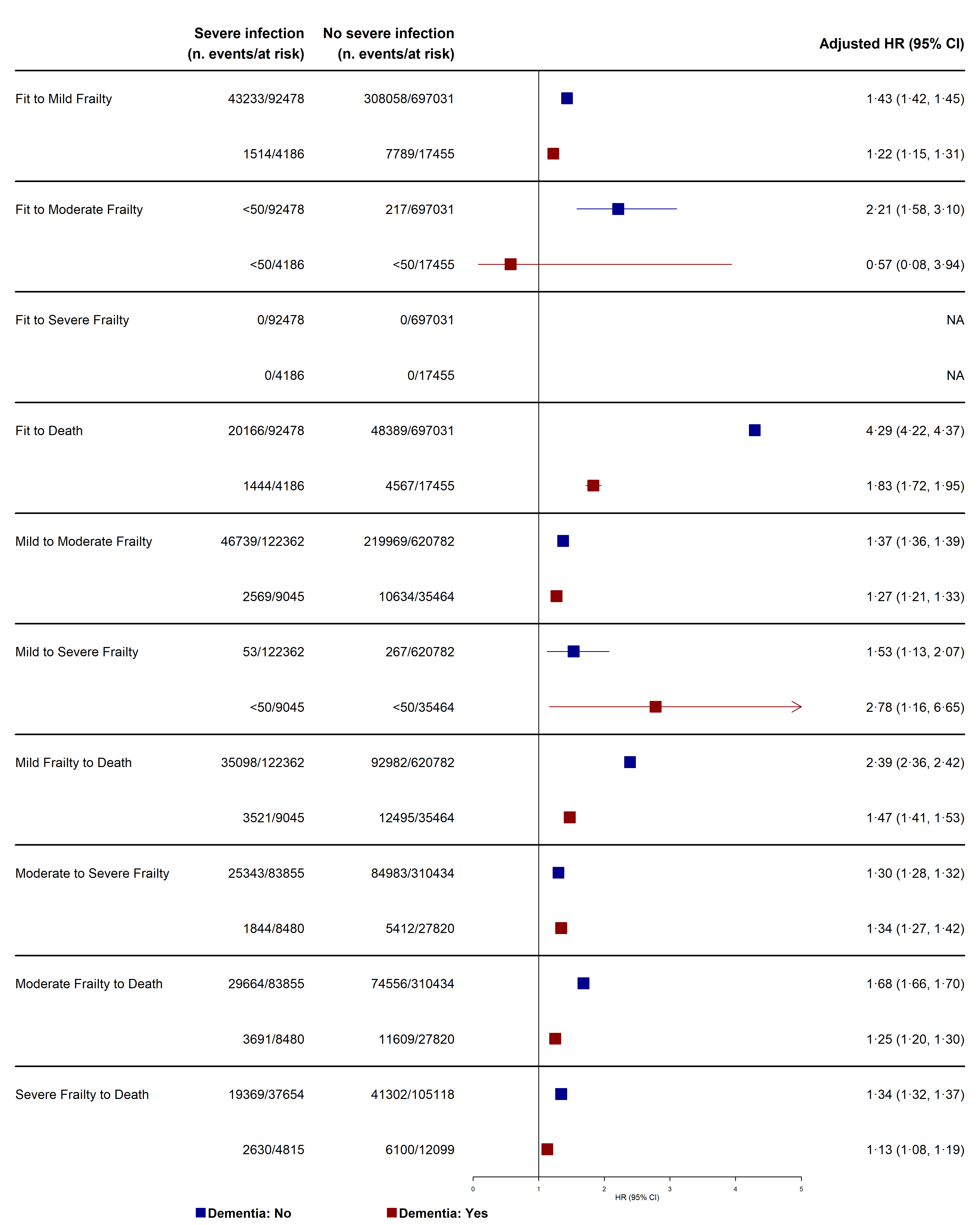

**Figure S10** Effect modification by dementia status. Forest plots of transition hazard ratios from the multistate model comparing adults with and without severe infection stratified by dementia status.

CI = Confidence Interval, HR = Hazard Ratio.

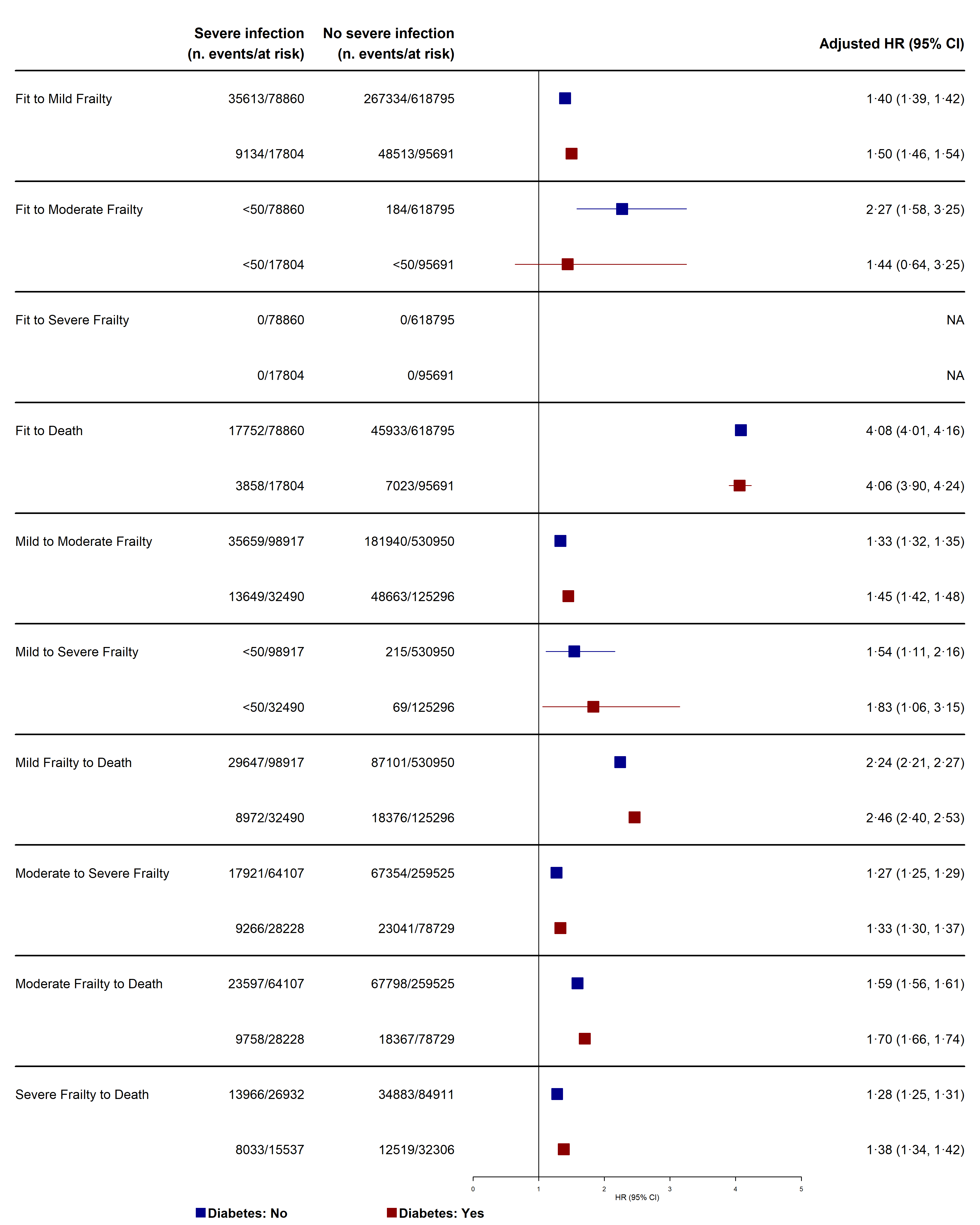

**Figure S11** Effect modification by diabetes mellitus status. Forest plots of transition hazard ratios from the multistate model comparing adults with and without severe infection stratified by diabetes mellitus status.

CI = Confidence Interval, HR = Hazard Ratio.

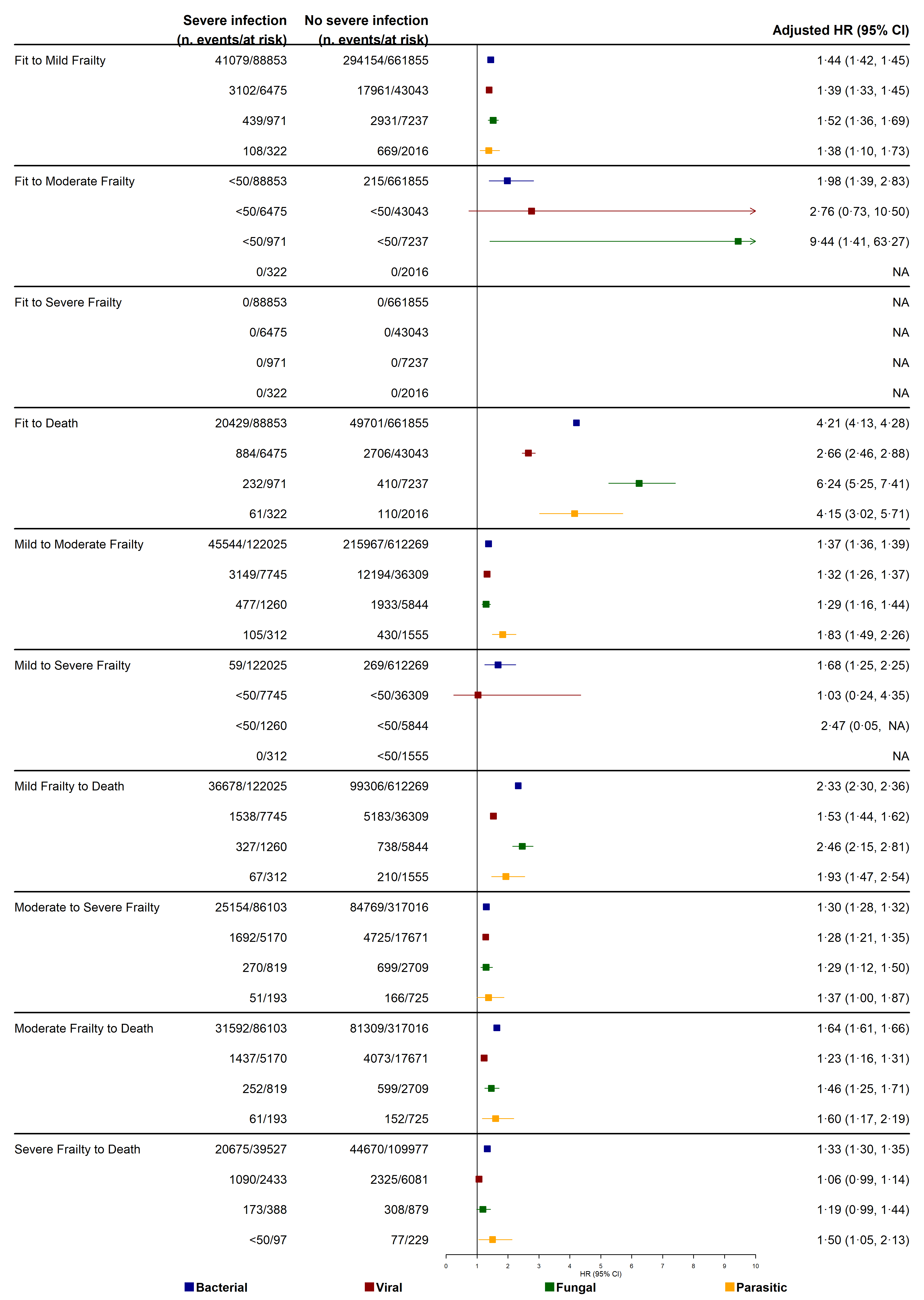

**Figure S12** Effect modification by infection pathogen. Forest plots of transition hazard ratios from the multistate model comparing adults with and without severe infection stratified by infection pathogen of the exposed individual, Bacterial, Viral, Fungal, and Parasitic.

CI = Confidence Interval, HR = Hazard Ratio.

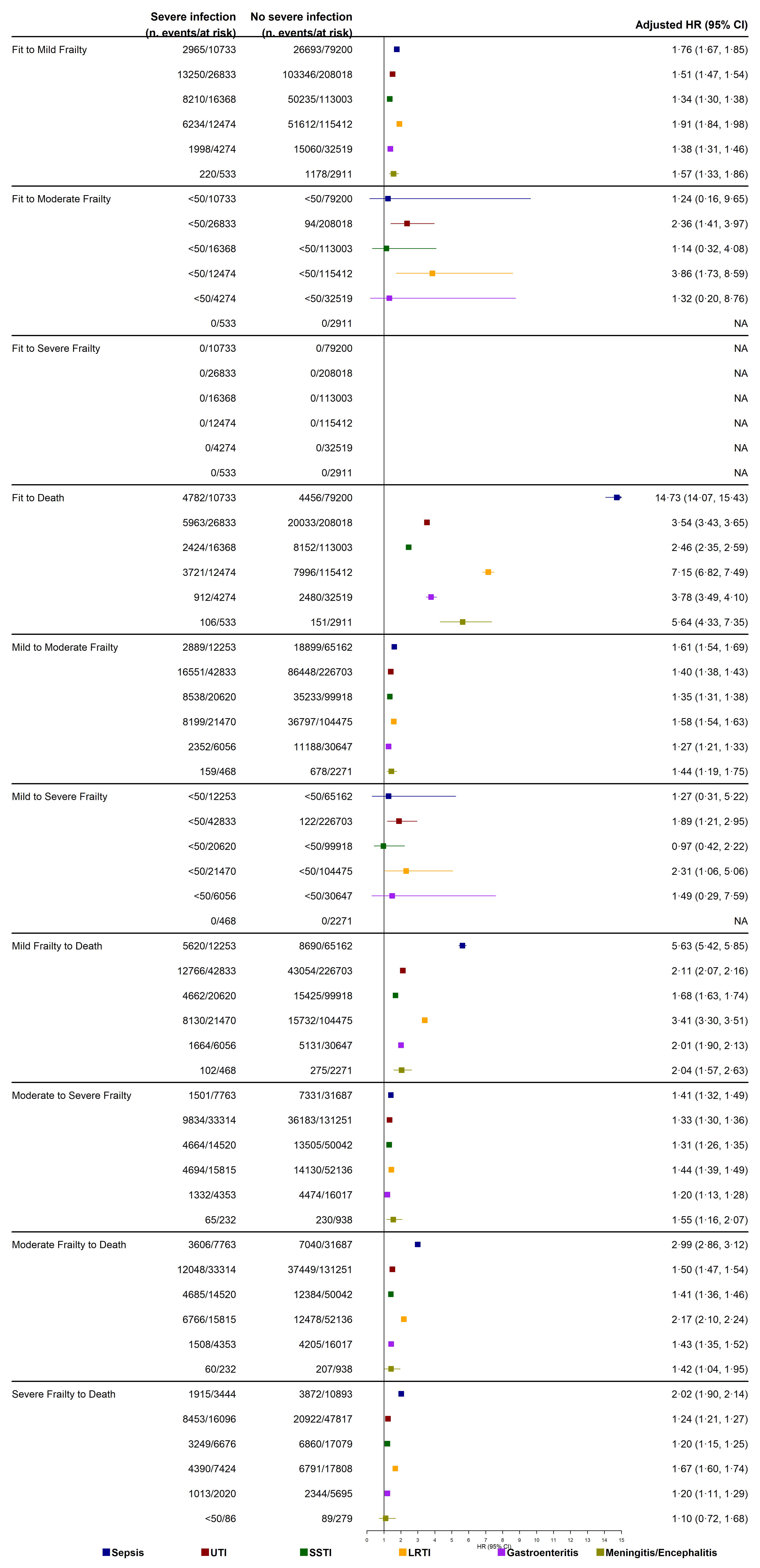

**Figure S13** Effect modification by infection type. Forest plots of transition hazard ratios from the multistate model comparing adults with and without severe infection stratified by infection type of the exposed individual: Sepsis, UTI, Skin and soft tissue, Gastroenteritis, LRTI, and Meningitis/Encephalitis.

CI = Confidence Interval, HR = Hazard Ratio, UTI=Urinary Tract Infection, LRTI = Lower Respiratory Tract Infection.
